# RT-LAMP assay for ultra-sensitive detection of SARS-CoV-2 in saliva and VTM clinical samples

**DOI:** 10.1101/2020.11.16.20232678

**Authors:** A. Ganguli, A. Mostafa, J. Berger, S. A. Stewart de Ramirez, A. Baltaji, K. Roth, M. Aamir, S. Aedma, M. Mady, P. Mahajan, S. Sathe, M. Johnson, K. White, J. Kumar, E. Valera, R. Bashir

## Abstract

The COVID-19 pandemic has underscored the shortcomings in the deployment of state-of-the-art diagnostic platforms. Although several PCR-based techniques have been rapidly developed to meet the growing testing needs, such techniques often need samples collected through a swab, the use of RNA extraction kits, and expensive thermocyclers in order to successfully perform the test. Isothermal amplification-based approaches have also been recently demonstrated for rapid SARS-CoV-2 detection by minimizing sample preparation while also reducing the instrumentation and reaction complexity. There are limited reports of saliva as the sample source and some of these indicate inferior sensitivity when comparing RT-LAMP with PCR-based techniques. In this paper, we demonstrate an improved sensitivity assay to test saliva using a 2-step RT-LAMP assay, where a short 10-minute RT step is performed with only B3 and BIP primers before the final reaction. We show that while the 1-step RT-LAMP demonstrate satisfactory results, the optimized 2-step approach allows for single molecule sensitivity per reaction and performs significantly better than the 1-step RT-LAMP and conventional 2-step RT-LAMP approaches with all primers included in the RT Step. Importantly, we demonstrate RNA extraction-free RT-LAMP based assays for detection of SARS-CoV-2 from VTM and saliva clinical samples.

---

The outbreak of the severe acute respiratory syndrome coronavirus 2 (SARS-CoV-2) pandemic has become a major challenge for national health care systems, infecting millions of people, burdening daily life, and causing heavy economic losses^1,2^. The fast spread of the COVID-19 pandemic has underlined the shortcomings of the existing technologies and testing paradigm for viral diagnostics and has propelled the need for alternate rapid and accurate diagnostic approaches for SARS-CoV-2 detection. While we await the SARS-CoV-2 vaccine, massive and repeated testing has been highlighted as a critical step to suppress the spread^3^. However, due to the current cost of the test, the time required to obtain results, and other logistical difficulties for massive deployment, there is further need for additional scientific improvements for the deployment of testing solutions.

The current gold standard approaches for viral detection primarily relies on polymerase chain reaction (PCR) but such assays often need extensive and time-consuming RNA purification steps and expensive thermocyclers to successfully perform the test. These assays are also challenging to perform at the point of care. Isothermal amplification techniques such Loop mediated isothermal amplification (LAMP)^4^ have generating much interest especially to eliminate the need for precise thermal cycles to achieve the nucleic acids amplification^5^. Likewise, LAMP has also shown to be robust against crude samples and inhibitors that often slow PCR^6,7^. In addition, its use of 4-6 primers to identify 6-8 regions of a target genome for amplification increases the specificity in comparison to PCR^4,8^. The robustness of the Bst polymerase and Reverse Transcriptase enzymes allow direct detection of RNA targets from crude samples without any RNA extraction or purification^6,9^. However, the sensitivity of these assays detecting RNA viruses has remained low and to the best of our knowledge never reached single molecule detection limits^10-12^. In the particular case of SARS-CoV-2, recent studies indicate that the sensitivity of RT-LAMP is inferior to that of some PCR-based techniques for COVID-19 in saliva specimens^13^.

Besides the detection technique, the sample specimen is also critical when developing assays that could be massively scaled or used for asymptomatic testing. Towards this end, although saliva is becoming a preferred and scalable option for reliable and massive testing, very few of the EUA approved PCR-based tests detect the SARS-CoV-2 RNA from saliva specimens^14^ and to the best of our knowledge to the date there are no saliva-to-RT-LAMP EUA approved tests. The use of saliva as sample has demonstrated not only to serve as an alternative upper respiratory tract specimen type for SARSCoV-2 detection^15-17^, but also saliva offers a number of advantages over nasopharyngeal (NP) and nasal swabs when considering the aforementioned criteria for mass testing efforts. The use of NP swabs can cause discomfort and irritation that could promote sneezing and coughing and increase the risk of exposure for the medical providers^18,19^. Collection of NP swabs has been associated with variable, inconsistent, and false negative test results^17^. On the other hand, saliva does not require a certified swab, specific collection receptacle, or transport media, and does not have to be obtained by a skilled healthcare provider, all of which increase diagnostic associated costs. Despite the interest generated and the mentioned advantages, only 3 papers in the pre-print form^20,21^ or peer-reviewed^13^ have been reported regarding RT-LAMP detection of SARS-CoV-2 RNA from saliva specimens. Although these papers highlight the possibility of bypassing the RNA extraction step^20,21^, they note the limitation on sensitivity of traditional RT-LAMP when detecting SARS-CoV-2 RNA from saliva. For instance, it is mentioned that RT-LAMP reaction was still not sufficiently sensitive to detect fewer than 200 viral copies/μL in saliva and that significant modifications to the RT-LAMP technique were needed to achieve the single-copy detection levels^20^. Also, an RT-LAMP colorimetric assay enabled detection of ∼ 100 viral genomes per reaction^21^, and in an study across 103 saliva specimens, the RT-LAMP show only 70.9% of sensitivity and its sensitivity was behind of some PCR-based techniques ^13^.

In this paper, we address the above limitations and present an optimized RT-LAMP approach for detecting viral RNA with single molecule sensitivity from NP swabs (Viral Transport Media) and saliva specimens. By adding a short 10-minute RT-incubation step with B3 and BIP primers specifically separated to improve primer annealing with target, we increase the sensitivity of the RT-LAMP reaction by 2 orders of magnitude over the current 1-step or 2-step RT-LAMP reactions. We first characterize our approach for detection of Zika virus as an example, and quantitatively demonstrate the improvement in limit of detection (LOD). We then use our optimized assay for detecting SARS-CoV-2 viral RNA and inactivated viruses in buffer to achieve single molecule sensitivity. Next, we show the robustness of our assay by doing direct detection of inactivated SARS-CoV-2 viruses in unprocessed VTM and saliva samples, bypassing the RNA extraction step, with a detection limit down to 1 copy/μL within 40 minutes, and with a sample to result time of less than 1 hour. Finally, we tested our RT-LAMP reactions detecting SARS-CoV-2 virus in 50 VTM and 34 saliva clinical samples. It is important to note that our protocol gives superior sensitivity to the conventional 2-step RT-LAMP protocol where reverse transcription (RT) is separately performed with all primers followed by an amplification step^22-24^. We believe, with our improved LOD and simple RNA extraction-free protocol, our approach will allow rapid scaling of testing and detection of cases which might have been otherwise missed due to low viral loads.

## Results

### Improved RT-LAMP assay

For sensitive detection of viruses, we optimized the standard 1-step RT-LAMP process flow to include a short incubation step for cDNA synthesis before the LAMP reaction. **Figure 1** compares our optimized 2-steps RT-LAMP protocol with the conventional 1-step RT-LAMP and 2-step RT-LAMP approaches. Our improved RT-LAMP assay is a 2-step process that begins with thermally lysed viral samples (95 °C, 10 minutes). We and others have recently shown that a short thermal lysis prior to RT-LAMP reaction efficiently exposes the RNA for amplification and is also known to inactivate nucleases in crude samples^5,25^. In our new protocol, post thermal lysis, we add LAMP buffer reagents, B3 and BIP LAMP primers, and the thermophilic Reverse Transcriptase enzyme (RTx, NEB), and perform a 10-minute RT incubation at 55 °C. The reduced temperature for the RT step (55 °C RT vs 65 °C LAMP) allows for improved annealing between the RNA and the primers and allows for efficient complementary strand synthesis. Since LAMP requires the formation of loop structures for amplification through a defined sequence of steps (binding and extension of inner primer, followed by binding and extension of outer primer), having only B3 and BIP in the RT step reduces the possibility of mis-priming events^26,27^ and allows for improved LOD which we characterize in the following sections. After the short RT incubation, the rest of the primers (F3, FIP, LF and LB) and Bst polymerase are added, and the LAMP reaction is performed at a constant temperature (65 °C, 60 min.). It is important to note that our modified protocol involving a separate RT step with only B3 and BIP primers offers superior sensitivity to the conventional 2-step RT-LAMP process where RT incubation is performed with either random hexamers or all of the LAMP primers^22-24^.

**Figure 1.**
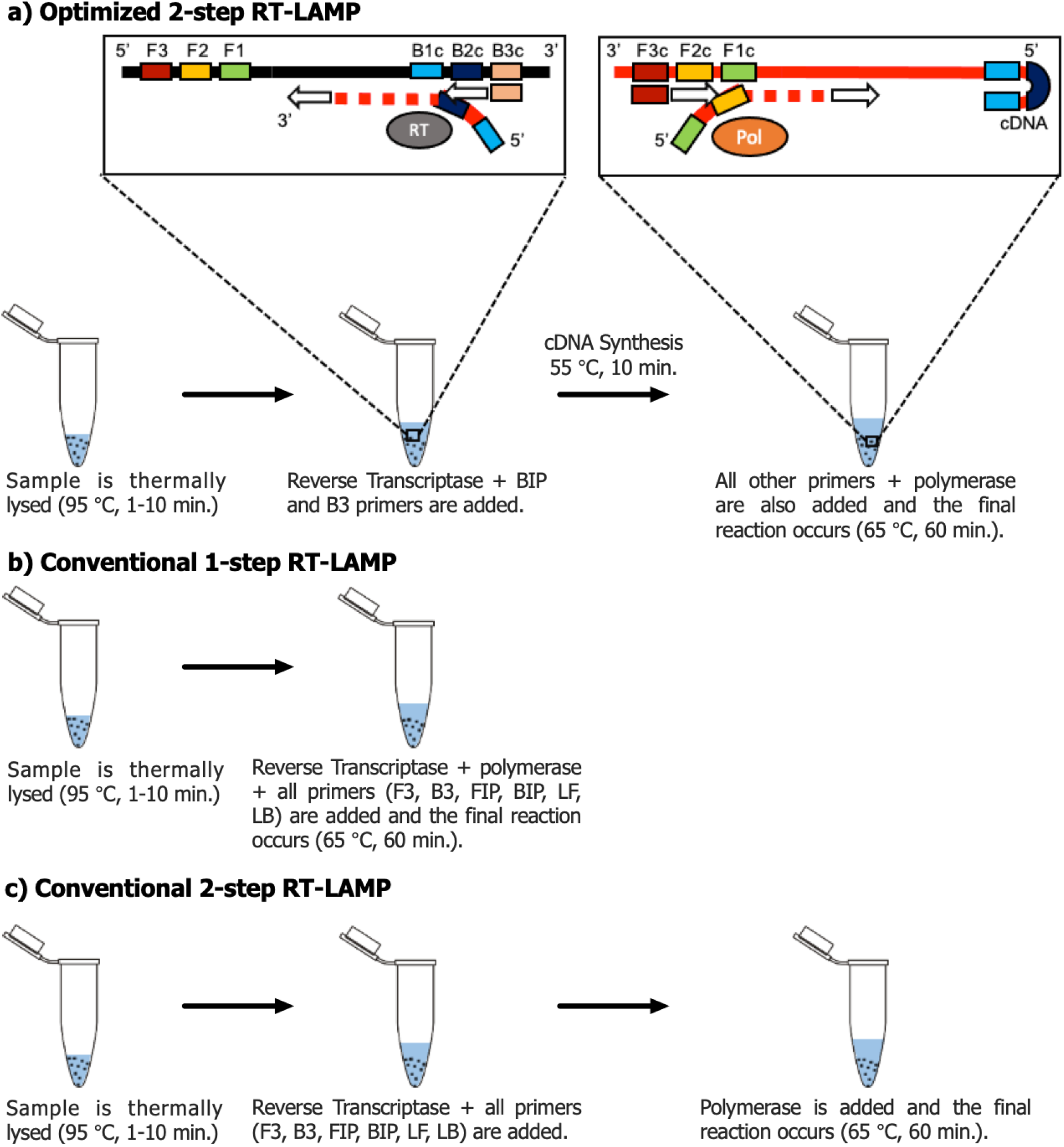
Process flow schematic of RT-LAMP approaches. **a)** Optimized 2-step RT-LAMP: Viral sample is first thermally lysed (95 °C, 1 min.). Then, RT-LAMP buffer reagents, B3 and BIP primers only, and reverse transcriptase enzyme is added. A short incubation (55 °C, 10 min.) is performed for cDNA synthesis to occur. Finally, the rest of the primers and Warmstart Bst 2.0 polymerase are added for the final reaction (65 °C, 60 min.); **b)** Conventional 1-step RT-LAMP: After thermal lysis, the RT-LAMP buffer reagents, polymerase and reverse transcriptase enzyme and all primers are added for the reaction to occurs (65 °C, 60 min.); **c)** Conventional 2-step RT-LAMP: After thermal lysis, the RT-LAMP buffer reagents, reverse transcriptase enzyme and all primers are added. Then, the polymerase is also added, and the reaction occurs (65 °C, 60 min.).

### Characterization with Zika RNA and virus

We characterized our optimized RT-LAMP assay by comparing the LODs of RT-LAMP assays with and without the short RT incubation step using Zika genomic RNA as target. **Figure S1a-c** (supplementary information) shows the amplification curves for the detection of Zika genomic RNA with: 1) our optimized 2-step reaction with a reverse transcriptase incubation (RT-incubation) step (55 °C, 10 minute), where only the B3 and BIP primers were added during the RT-incubation (all other primers were added for the final reaction at 65 °C) (**Figure S1a**); 2) in a 1-step RT-LAMP reaction (no intermediate incubation step) (**Figure S1b**); and 3) in a 2-step reaction process with an intermediate RT-incubation at 55 °C (10 min.) and where all primers were added (**Figure S1c**). **Figure S1d** shows the threshold times plot for these reactions. While in the case of the 1-step RT-LAMP reaction the LOD was 2×10^3^ copies/μL, in the traditional 2-step RT-LAMP reaction the LOD improved to 2×10^2^ copies/μL with 2/3 amplifications for 2×10^1^ copies/μL. However, when using our optimized 2-step RT-LAMP reaction, the LOD was as low as 2 copies/μL (2/3 replicates were amplified). This improved LOD is 3 orders of magnitude better than the detection limit of a 1-step RT-LAMP reaction. Note that the limit of detection is determined by the expected sampling frequency and as we approach single molecules, sampling bias can start to occur.

Then, we evaluated the efficiency of our optimized 2-step RT-LAMP assay to directly detect viruses using Zika virus as the target. We serially diluted Zika viruses and spiked them in our reactions. **Figure S1e-f** shows the amplification curves for the detection of Zika virus in a 1-step RT-LAMP reaction and in our optimized 2-step RT-LAMP reaction. While in the 1-step RT-LAMP reaction the LOD was 4×10^2^ copies/μL (only 1/3 amplifications for 4×10^1^ copies/μL), in the case of our optimized reaction the obtained LOD was 4×10^1^ copies/μL with all three replicates amplifying in less than 20 min. **Figure S1g** shows the threshold times plot for these reactions. Though the detection limit of Zika virus did not reach 4 copies/μL, we observed 1 order of magnitude improvement in detection limit with our 2-step RT-LAMP process as well as faster amplification timings for the 4×10^3^ and 4×10^2^ copies/μL target concentrations.

### Characterization with SARS-CoV-2 RNA and virus

We next evaluated our assay composition and protocol with SARS-CoV-2 genomic RNA and inactivated viruses. Thus, we first confirmed the improved detection limit of our optimized RT-LAMP assay by performing the 1-step and our modified 2-step reactions to detect SARS-CoV-2 genomic RNA. **Figure S2a-b** shows the amplification curves for the detection of SARS-CoV-2 genomic RNA in a 1-step RT-LAMP reaction and in our optimized 2-step RT-LAMP reaction. While in the 1-step RT-LAMP reaction the LOD was 2×10^1^ copies/μL, we observed that our optimized 2-step assay allowed an improved LOD of 2 copies/μL with all three replicates amplifying within 25 minutes. **Figure S2c** shows the threshold times plot for these reactions. These results highlight that our optimized RT-LAMP assay is able to amplify 2 copies/μL of SARS-CoV-2 RNA in starting sample.

Next, we compared the results between a 1-step RT-LAMP assay and our optimized 2-step assay for the detection of inactivated SARS-CoV-2 virus. The LOD was 1 copy/μL using our optimized protocol with a short RT-incubation and separated primers (**Figure S2d-e**). This is more than 1 order of magnitude sensitive than the LOD of the 1-step RT-LAMP assay for SARS-CoV-2 viruses. **Figure S2f** shows the threshold times plot for these reactions We further observed 3-5 minutes faster amplification timings for the three highest concentrations of viral sample (4×10^3^-4×10^1^ copies/μL) in the 2-step RT-LAMP process. These results highlight that our optimized RT-LAMP assay can show single molecule sensitivity.

### Detection of SARS-CoV-2 virus in Viral Transport Media

Current diagnostic testing for SARS-CoV-2 includes collecting nasopharyngeal or anterior nasal specimen swabs with viruses and transferring them into viral transport media (VTM), from which the sample can be collected, purified, and tested using a diagnostic assay^28^. To evaluate if the detection limit of viral samples in VTM is improved by our optimized 2-step RT-LAMP reaction, we spiked serial dilutions of inactivated SARS-CoV-2 virus in VTM and performed both the usual 1-step and our optimized 2-step RT-LAMP reactions. The amplification curves (**Figure 2a-b**) and threshold times (**Figure 2c**) show a detection limit of 1 copy/μL for our optimized RT-LAMP assay in comparison to the LOD of 40 copies/μL in the 1-step RT-LAMP process. These results demonstrate no reduction in sensitivity of our assay in VTM compared to viruses spiked in buffer. It also highlights the fact that for our assay sensitive detection is possible without any RNA extraction. This pathway can allow potential integration in the current clinical workflow from nasopharyngeal or nasal samples.

**Figure 2.**
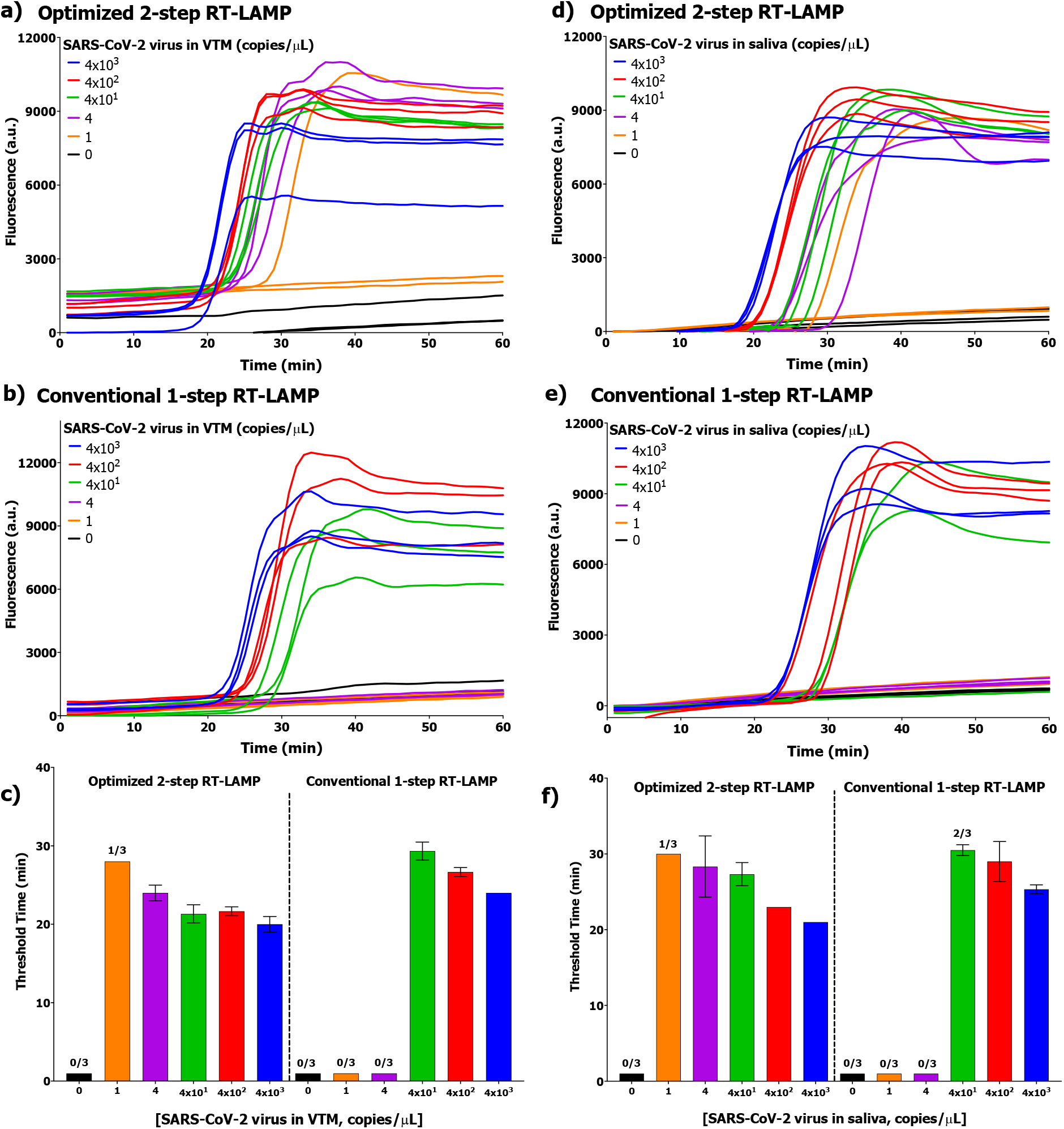
Detection of inactivated SARS-CoV-2 virus in VTM and human saliva. **a-b)** Raw amplification fluorescence curves for inactivated SARS-CoV-2 virus detection in VTM. **c)** Threshold timings for inactivated SARS-CoV-2 virus in VTM. Optimized 2-step RT-LAMP LOD = 1 copies/μL; Conventional 1-step RT-LAMP LOD = 4×10^1^ copies/μL. **d-e)** Raw amplification fluorescence curves for inactivated SARS-CoV-2 virus detection in saliva; **f)** Threshold timings for inactivated SARS-CoV-2 virus detection in saliva. Optimized 2-step RT-LAMP LOD = 1 copies/μL; Conventional 1-step RT-LAMP LOD = 4×10^2^ copies/μL. The bar graphs show mean and standard deviation for 3 replicates.

### Detection of SARS-CoV-2 virus in spiked healthy human saliva

The use of saliva has been demonstrated as an alternative upper respiratory tract specimen type for SARS-CoV-2 detection and also to offer advantages over the use of NP and nasal swabs. Thus, to evaluate our optimized assay with unprocessed saliva samples, serial dilutions of inactivated SARS-CoV-2 viruses were spiked in purchases pooled human saliva (Innovative Research). The detection was performed using the conventional 1-step as well as our optimized 2-step RT-LAMP reaction. In the amplification curves (**Figure 2d-e**) and threshold times (**Figure 2f**), we observed that for the 1-step RT-LAMP reaction, a LOD of 4×10^2^ copies/μL virus in saliva was obtained (with 2/3 replicates amplifying for 4×10^1^ copies/μL). In contrast, in our optimized 2-step RT-LAMP assay in saliva, all three replicates amplified for 4 copies/μL and 1/3 replicates amplified for 1 copy/μL. This highlights the single molecule sensitivity with which we can detect SARS-CoV-2 viruses from unprocessed human saliva samples without any purification or extraction of RNA. These results were again confirmed when repeating the experiments under the same conditions (**Figure S3**).

### Detection of SARS-CoV-2 virus from clinical samples

In order to demonstrate the applicability of our assay, we also characterized our optimized 2-step RT-LAMP reaction using 50 VTM clinical samples (25 known positives and 25 known negatives) and 34 saliva clinical samples (from in-patients at Carle Foundation Hospital). While for the analysis of the VTM samples we used our optimized 2-step RT-LAMP reaction (2 μL sample + 14 μL reaction mix), for the characterization of our assay with saliva samples we compared our 2-step RT-LAMP reaction with the conventional 1-step RT-LAMP reaction and also using two different reaction volumes to examine differences in sampling. For saliva, we used the small volume format (2 μL sample + 14 μL reaction mix) and also the large volume format (12 μL sample + 84 μL reaction mix) for both optimized 2-step and traditional 1-step reactions. Thus, in the case of saliva, four different reactions were used to analyze each of clinical samples. In addition to the presence of the SARS-CoV-2 virus, the quality of samples (both VTM and saliva) was also tested by spiking an aliquot of the sample with MS2 bacteriophage (internal control). **Figure 3** summarize the samples, techniques, and volume formats used. Each clinical sample (both VTM and saliva) was also tested using the RT-PCR assays from clinical lab results (gold standard), which included the RNA purification step, and were used to compare our RT-LAMP results.

**Figure 3.**
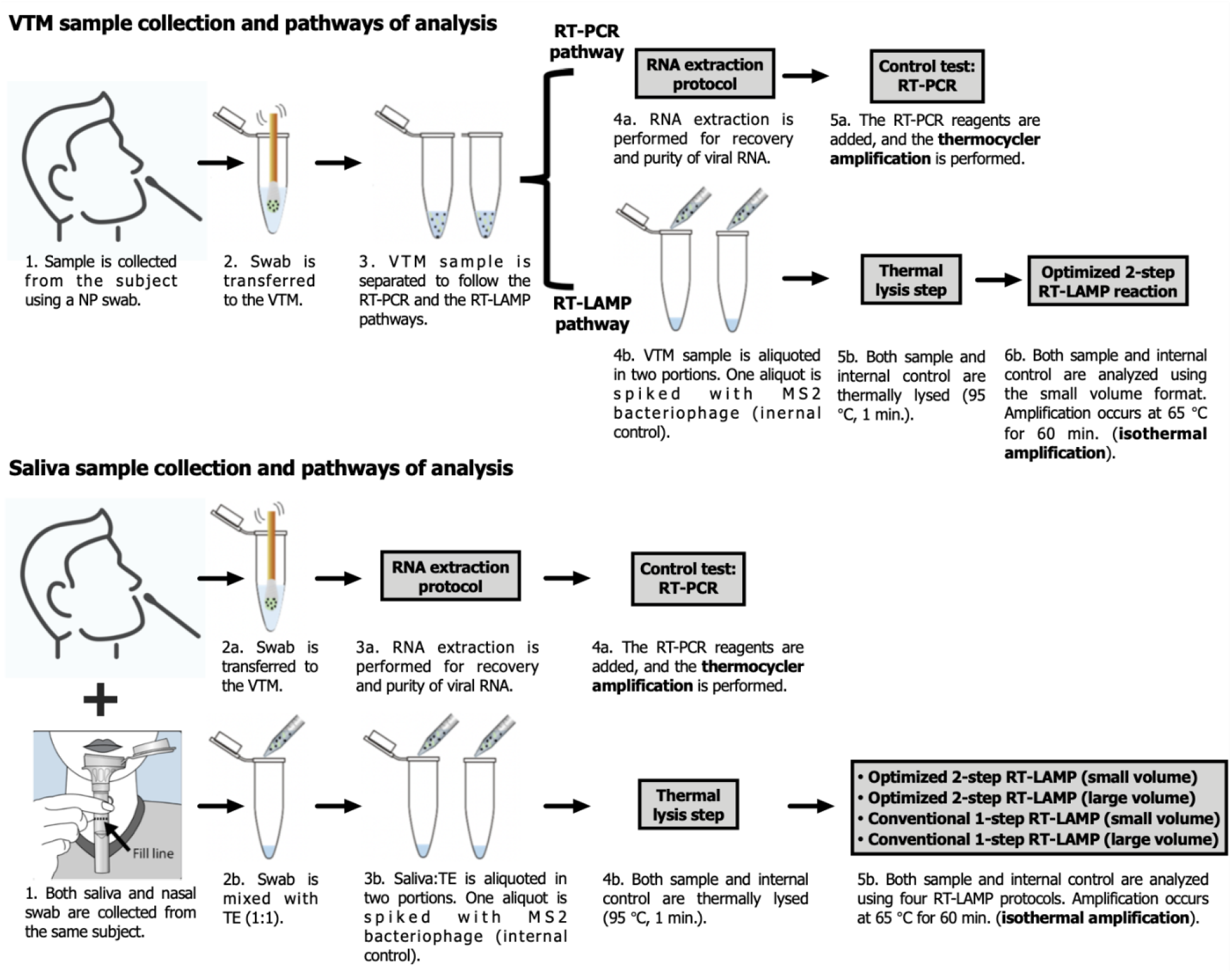
VTM and saliva samples collection and pathways of analysis. VTM samples (NP swab collection) were analyzed using our optimized 2-step RT-LAMP reaction (2 μL sample + 14 μL reaction mix) and RT-PCR gold standard technique (control). Saliva samples and nasal swab (collected from the same subject) were analyzed using RT-LAMP and RT-PCR (control) techniques respectively. For the RT-LAMP technique we compared our 2-step RT-LAMP reaction with the conventional 1-step RT-LAMP reaction using the small volume (2 μL sample + 14 μL reaction mix) and the large volume (12 μL sample + 84 μL reaction mix) formats. In the case of RT-LAMP techniques, for both VTM and saliva, the quality of samples was also tested by spiking an aliquot of the sample with MS2 bacteriophage (internal control).

The VTM samples used in our work were obtained from Order of St. Francis (OSF) Healthcare (Peoria, IL) through an approved institutional review board (OSF Peoria IRB # 1602513 through the University of Illinois College of Medicine at Peoria with waiver for consent). The samples received were VTM discards prior to the RNA purification step. Along with the samples, we also received the results of the RT-PCR tests performed by OSF Healthcare. The results obtained using these VTM clinical samples are summarized in **Figure 4** (raw amplification data is in **Figure S4**). Our RNA extraction-free 2-step RT-LAMP reaction (small volume) was able to detect as positives, 23 of 25 confirmed VTM positive samples, after a thermal lysis step (95 °C, 1 minutes). Likewise, this reaction was able to detect as negatives, 25 of 25 confirmed VTM negative clinical samples. With these results, the Positive Predictive Value (PPV), Negative Predictive Value (NPV), sensitivity, and the specificity were calculated as 100%, 92.6%, 92%, and 100% respectively.

**Figure 4.**
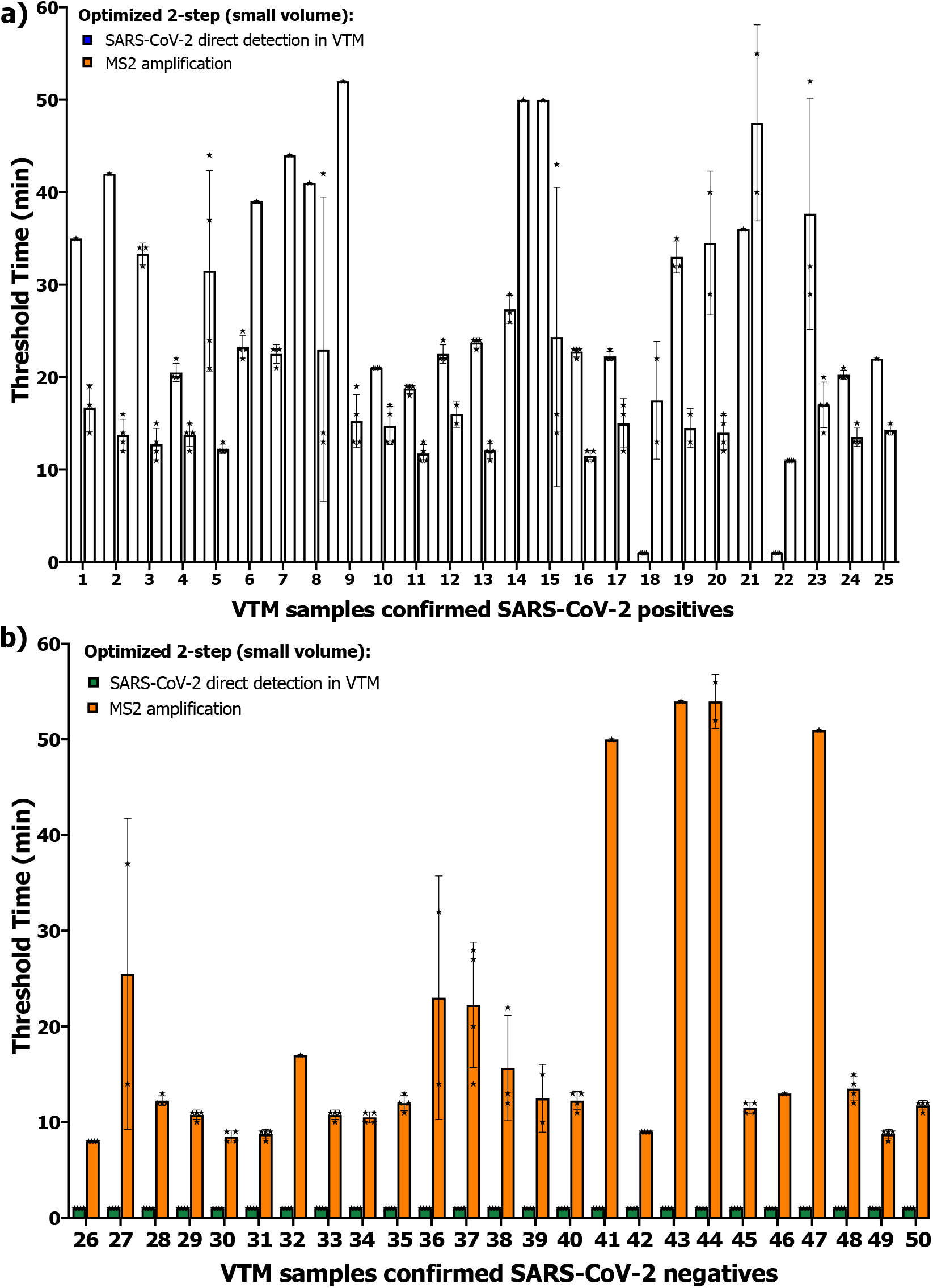
Summary of the results obtained when analyzing VTM clinical samples with our 2-step RT-LAMP reaction (small volume). Samples used were VTM discards prior to the RNA purification step. **a)** Confirmed positive samples (n = 4). Our reaction detected as positives, 23 of 25 confirmed VTM positive samples. **b)** Confirmed negative samples (n = 4). Our reaction detected as negatives, 25 of 25 confirmed VTM negative samples. For all positive and negatives samples, internal control was also analyzed (n = 4). Symbols in the bars indicate the results of the individual replicates.

The saliva samples used in this work were obtained from COVID-19 in-patients at Carle Foundation Hospital (Urbana, IL.) through an approved institutional review board (Carle IRB # 20CRU3150). Along with the collection of the saliva clinical samples, nasal swab samples were also collected from the same subject at Carle Foundation Hospital and analyzed by RT-PCR technique at Carle clinical lab. Immediately after collection, the saliva samples were mixed with TE buffer (1:1). The use of TE buffer reduced the viscosity of the sample, allowing for easier pipetting. The results obtained using these saliva clinical samples are summarized in **Figure 5** (amplification raw data and internal control results are in **Figure S5**).

**Figure 5.**
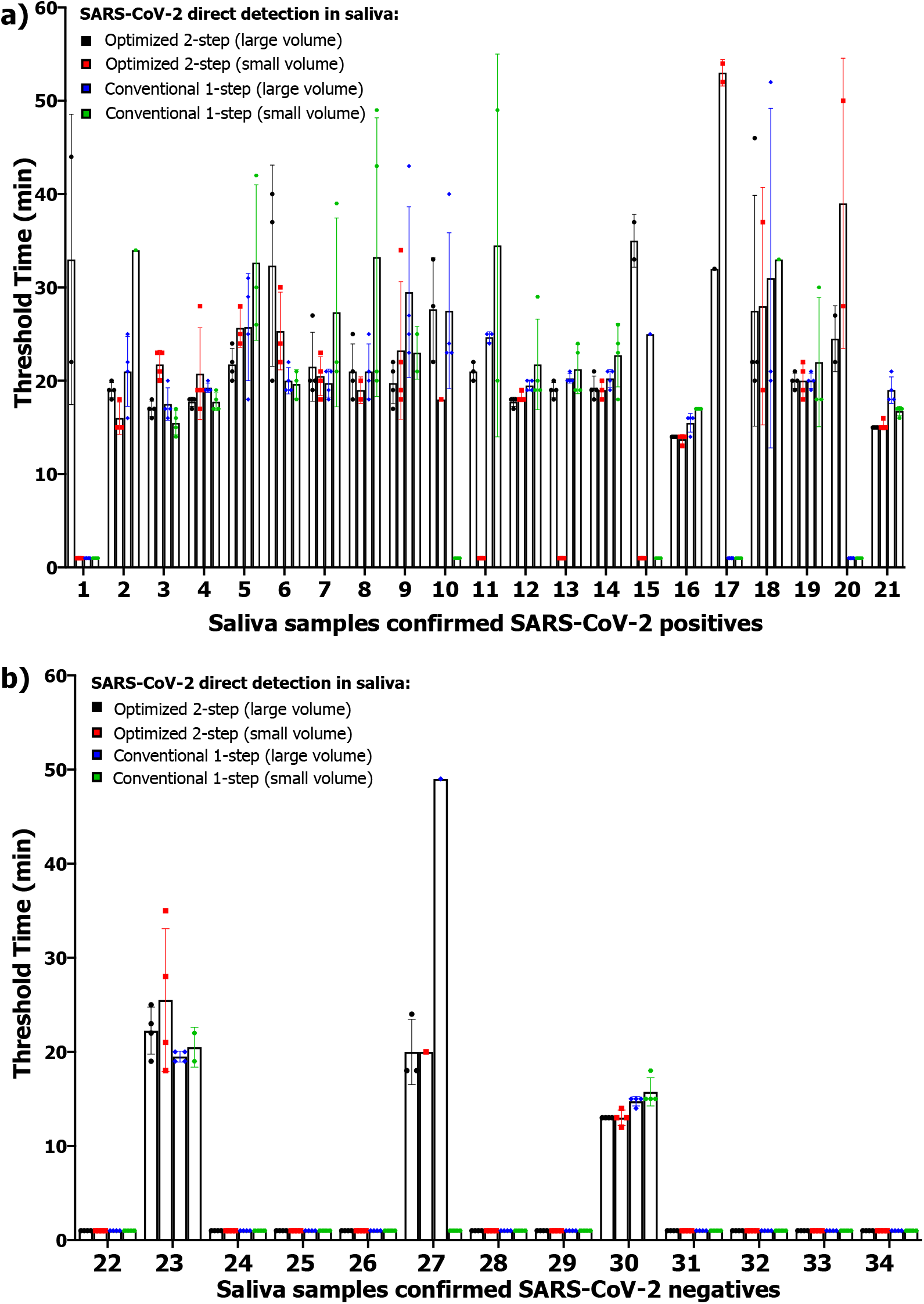
Summary of the results obtained when analyzing saliva clinical samples. Samples used were saliva from in-patients for COVID-19. **a)** Confirmed positive samples analyzed with our optimized 2-step RT-LAMP reaction (large and small volume) and also with conventional 1-step RT-LAMP reaction (large and small volume) (n = 4). **b)** Confirmed negative samples analyzed with our optimized 2-step RT-LAMP reaction (large and small volume) and also with conventional 1-step RT-LAMP reaction (large and small volume) (n = 4). Symbols in the bars indicate the results of the individual replicates.

Results demonstrate that our 2-step RT-LAMP reaction (large volume) offered the best results with 100% of sensitivity when analyzing the 21 saliva samples that were confirmed positive with the RT-PCR results from clinical laboratory from the nasal swab control samples. Interestingly, from the 13 saliva samples confirmed negative, 3 where detected as positive with our method. Note that samples (both confirmed positive and negative) are from patients admitted in the hospital who were diagnosed COVID-19 positive by RT-PCR of nasal samples obtained within 14 days of the saliva sample collection date.

## Discussion

Although nasopharyngeal, oropharyngeal, and nasal swabs have been recommended by the CDC as the upper respiratory tract specimen types for SARS-CoV-2 viral testing^28^, saliva specimens have been an appealing alternative to the swabs, as saliva samples can be collected non-invasively, and minimizes the contact between healthcare workers and patients and the chance of exposure to the virus^16,17^. In previous analysis of the concordance between nasopharyngeal and saliva samples for detection of SARS-CoV-2 and other respiratory pathogens, it has been demonstrated that sensitivity between the two sample types are highly comparable^16,17,29,30^. Therefore, in this paper, we evaluated the detection of SARS-CoV-2 virus without RNA extraction not only from VTM clinical samples but also from saliva clinical samples using the optimized 2-step RT-LAMP protocol in the current clinical workflow. In addition, in the case of saliva, we studied the role of the sampling volume in the assay sensitivity. For saliva clinical samples, we used the small volume format (2 μL sample + 14 μL reaction mix) and also the large volume format (12 μL sample + 84 μL reaction mix) for both optimized 2-step and conventional 1-step RT-LAMP reactions.

The VTM patient samples used in this work were collected and stored frozen prior to the RNA purification step at OSF Hospital (25 confirmed positive and 25 confirmed negative). The saliva samples used in this work were obtained from in-patients for COVID-19 (21 confirmed positive and 13 confirmed negative). In both cases, the presence of SARS-CoV-2 virus was confirmed using RT-PCR technique from NP (for VTM) and nasal swabs (for Saliva) as the control. First, we tested the VTM samples using our optimized 2-step RT-LAMP approach with the small volume format (2 μL sample + 14 μL reaction mix) due to limited sample amount available. The results (**Figure 4**) indicated a perfect agreement with the RT-PCR control when testing the negative samples. However, from the 25 positive samples, 2 were detected as negatives with our reaction in the small volume format. We believe this could be attributed to the fact that the concentration in these samples was low and the RNA molecules was not sampled in the volume that we analyzed. Results for PPV, NPV, sensitivity, and specificity were calculated based on these results (VTM in **Figure 6a** and **Figure 6b**). As shown in **Figure 6a-b**, our optimized 2-step RT-LAMP approach (small volume) show 100% PPV and specificity. However, due to the 2 false negatives, NPV and sensitivity were 92.6% and 92% respectively. Based on these results we decided to study the role of the sampling in the assay sensitivity by including a large volume format when analyzing saliva samples.

**Figure 6.**
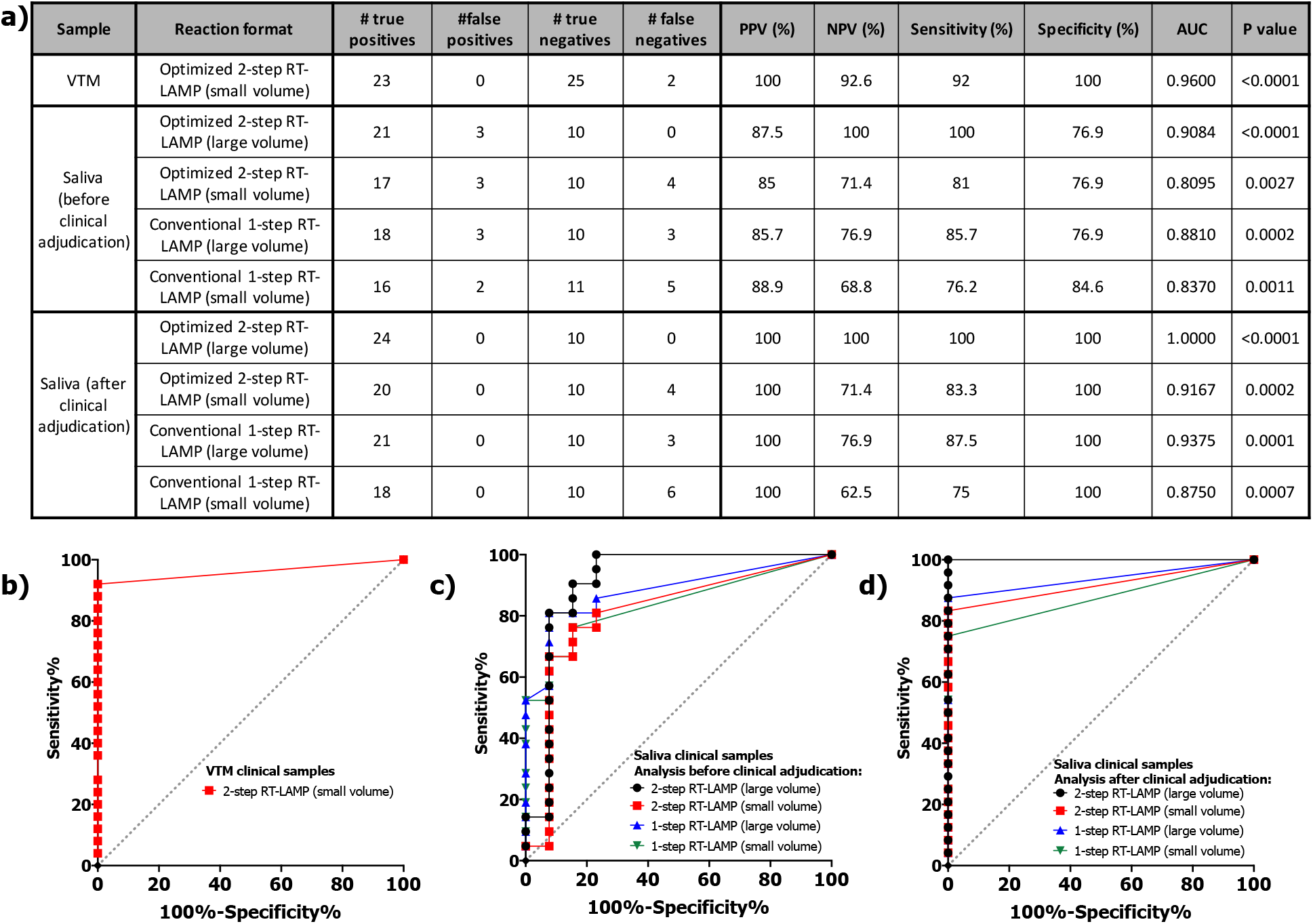
Summary of the performance of the different reaction conditions when analyzing VTM and saliva clinical samples. **a)** Summary table including results for VTM and saliva (before and after the clinical adjudication about samples 23, 27, and 30). **b-d)** ROC curves analysis for **b)** VTM clinical samples, **c)** saliva clinical samples (before clinical adjudication), and **d)** saliva clinical samples (after clinical adjudication). For each reaction condition, the positive samples were analyzed against the negative samples.

We found that when using a larger saliva sample volume in the optimized 2-step RT-LAMP approach, we obtained 100% sensitivity as we were able to detect as positives the 21 confirmed positives samples (**Figure 5a**, saliva - before clinical adjudication - in **Figure 6a**, and **Figure 6c**). However, 3 of the confirmed negative samples (# 23, 27, and 30) from the in-patients were detected as positives with our reaction impacting the specificity results (76.9%) (**Figure 5b**). Similar behavior was obtained when using the optimized 2-step RT-LAMP approach (small volume) and conventional 1-step RT-LAMP approach (large volume) where samples 23, 27, and 30 were also detected as positives. However, in these cases the sensitivity was worse as these reactions reported 4 and 3 false negatives, respectively. Finally, the conventional 1-step RT-LAMP approach (small volume) reported the lowest sensitivity (76.2%) although the specificity was a little bit better (84.6%) as only sample numbers 23 and 30 (not sample 27 in this case) were detected as positives. Interestingly, since the patients were known to be COVID-19 positive from an earlier test when admitted to the hospital due to symptoms, and were still in the hospital being treated when the saliva sample and the control nasal swab sample was taken, it could be hypothesized that the saliva samples depict the real presence of the viral RNA and the nasal swab does not. If samples 23, 27, and 30 are considered positives (after clinical adjudication), then sensitivity and specificity change as shown in **Figure 6a**, and **Figure 6d**. First, the total number of positives is now 24, while the total number of negatives is now 10. The optimized 2-step RT-LAMP approach (large volume) indicates a perfect agreement with the control (PPV, NPV, sensitivity, and specificity = 100%). In addition, for all reaction conditions studied, the specificity would be 100%. We can conclude that increasing the sample volume, improve the reaction sensitivity, although this also increase the cost of the assay due to increase use in reagents. Also, the use of the optimized 2-step approach improves the assay sensitivity in comparison with the conventional 1-step reaction when the same volume of sample is used. An important point to note is that the saliva samples can detect the presence of RNA in in-patient samples diagnosed of COVID-19 when the nasal swabs were not.

The better performance of the optimized 2-step RT-LAMP reaction (large volume) is also highlighted in the Receiver Operating Characteristic (ROC) curves (**Figure 6c-d**). With this reaction condition (after clinical adjudication) the AUC was 1.00, showing that this reaction can correctly detect positive and negative samples without false positives or false negatives. The saliva results and the study of the volume sampling indicate that the optimized 2-step RT-LAMP reaction (small volume) can produce some false negatives. This could explain the 2 false negatives we obtained when analyzing the VTM clinical samples.

It is important to highlight that although the sensitivity can vary with the different approaches used in this study, the RT-LAMP reaction achieves 100% specificity when analyzing VTM and saliva samples, independent of the assay conditions. This improved performance along with the high sensitivity of our optimized 2-step RT-LAMP approach are very important features when considering the use of RT-LAMP reactions for massive surveillance testing especially for portable and point of care applications^5^.

## Methods

### Zika and SARS-CoV-2 Genomic RNA and Viruses

We obtained ZIKV (PRVABC59) genomic RNA, NR-50244, and SARS-CoV-2 (Isolate USA-WA1/2020) genomic RNA, NR-52285, from BEI Resources and stored them in -80 °C. Stock volumes of genomic RNA were diluted to the correct concentrations in water. We also obtained whole viruses from BEI resources for ZIKV (PRVABC59), NR-50240, as well as heat-inactivated SARS-CoV-2, NR-52286. The stock vials were aliquoted and stored at -80 °C until used for experimentation. To obtain the appropriate concentrations, viruses were diluted in Tris-EDTA Buffer or VTM.

### Primer Sequences and Primer Validation Reactions

The primer sequences for the Zika and SARS-CoV-2 RT-LAMP reactions used in this paper were obtained from our previously published work^5,9^ and were synthesized by Integrated DNA Technologies.

### Clinical samples

VTM clinical samples used are discarded viral transport medium prior to the RNA purification step from 25 samples from patients who were tested positive for COVID-19 and from 25 samples from patients who were tested negative for COVID-19 at OSF Healthcare (Peoria, IL) by a RT-PCR test performed at OSF Healthcare. The samples were received de-identified, frozen and were obtained through an approved institutional review board (OSF Peoria IRB # 1602513 through the University of Illinois College of Medicine with waiver for consent).

Saliva clinical samples were collected from 34 in-patients at Carle Foundation Hospital (Urbana, IL) who were tested COVID-19 positive (RT-PCR) when they were admitted in the hospital (or at most 14 days before admission) through an approved institutional review board (Carle IRB # 20CRU3150). Simultaneously to the collection of the saliva clinical samples, nasal swab samples were also collected from the same subject at Carle Foundation Hospital and analyzed by RT-PCR technique at Carle clinical lab. Immediately after collection, the saliva samples were mixed with TE buffer (1:1). Samples were received de-identified and either fresh (during the collection day) or frozen (the next day after the collection day).

### RT-LAMP Reactions

The following components comprised the RT-LAMP assay: 4 mM of MgSO4 (New England Biolabs), 1× final concentration of the isothermal amplification buffer (New England Biolabs), 1.025 mM each of deoxyribonucleoside triphosphates, and 0.29 M Betaine (Sigma-Aldrich). Individual stock components were stored according to the manufacturer’s instructions, and a final mix including all of the components was freshly created prior to each reaction. Along with the buffer components, a primer mix consisting of 0.15 μM F3 and B3, 1.17 μM forward Inner primer (FIP) and backward inner primer (BIP), and 0.59 μM of LoopF and LoopB was added to the reaction. Finally, 0.47 U/μL BST 2.0 WarmStart DNA Polymerase (New England Bioloabs), 0.3 U/μL WarmStart Reverse Transcriptase (New England Biolabs), 1 mg/mL bovine serum albumin (New England Biolabs), and 0.735× EvaGreen (Biotium) were included in the reaction. EvaGreen dye is a double-stranded DNA intercalating dye. After addition of the template, the final volume of the reaction was 16 μL (small volume format) or 96 μL (large volume format). In the case of the small volume format, the volume of the template/sample was 2 μL, while in the case of the large volume format, the volume of the template/sample was 12 μL.

All RT-LAMP assays were carried out in 0.2 mL PCR tubes in an Eppendorf Mastercycler realplex Real-Time PCR System at 65 °C (60 min). Fluorescence data were recorded every 1 min. after each cycle of the reaction. While all characterization experiments were performed with number of replicates n = 3, n = 4 was used for the analysis of clinical samples.

For the 1-step RT-LAMP reactions without incubation, the first step was to serially dilute genomic RNA or viruses in H_2_O and aliquot 2 or 12 μL sample of the correct concentration into PCR tubes. Samples with viruses were thermally lysed (95 °C, 1 min.). Finally, 14 or 84 μL of RT-LAMP reagents were added with final concentrations as mentioned above, and the final reactions were incubated in the thermocycler for amplification.

For the 2-step RT-LAMP reactions with RT-incubation, the first step was to perform serial dilutions of the RNA or viral sample as mentioned in the 1-step protocol and to aliquot 2 or 12 μL sample of the correct concentration into PCR tubes. Only samples spiked with viruses were lysed (95°C, 1 min.) in a heater. Post thermal lysis, we added 3.28 μL (small volume format) of a buffer mix with B3 and BIP primers as well as reverse transcriptase enzyme and incubated the reaction tubes in a heater for 10 minutes at 55 °C (RT-incubation step). In this step, BIP and B3 primers anneal to the target and begin cDNA synthesis, without competition from other primers, reducing primer dimers. Post RT-incubation, 10.71 μL (small volume format) of the rest of the reagents for RT-LAMP were mixed in the reaction tubes and final reactions were incubated in the thermocycler for amplification. Final concentrations of all reagents in the two-step RT-LAMP assay were the same as mentioned above.

All RT-LAMP reactions consisted were performed with at least one set of non-template negative controls.

### SARS-CoV-2 Genomic RNA and Virus Detection in RT-LAMP Reactions

Reactions done with heat-inactivated viruses and clinical samples included a thermal lysis step. Heat-inactivated viruses were serially diluted in TE Buffer and then thermally lysed in a heater at 95 °C (1 min) prior to their addition into the final reaction mix. VTM clinical samples were thermally lysed in a heater at 95 °C (1 min) prior to their addition into the final reaction mix, while in the case of saliva clinical samples the thermal lysis step lasted 10 min. After heat lysis, the tubes were centrifuged (2000 g, 1 min.) and then kept in 4 °C (5 min.).

In the first analysis of VTM clinical samples we noticed that samples # 15, 18, 22, and 25 did not show amplification. Likewise, internal controls from samples # 21, 24, and 25 did not show amplification either. During processing we noticed that these samples were highly viscous and contained debris, which may have prevented amplification. We repeated the analysis of these samples but added a 5 min centrifugation step (4000 g). This additional centrifugation allowed separation of the debris and viscous components from the viral particles. The amplification reaction these samples was repeated using the supernatant post centrifugation. After the second analysis, internal controls from samples # 21, 24, and 25 did show amplification while from the VTM clinical samples only samples # 15 and 25 did show amplification. Results in **Figure 4, Figure 6a-b**, and **Figure S4** report the second analysis of these samples.

VTM used for assay characterization was obtained from Redoxica (VTM-500ML), aliquoted, and stored in 4 °C away from direct light. Saliva used for assay characterization was pooled human saliva obtained from Innovative Research (IRHUSL5ML) and aliquoted and stored in -20 °C until use for the reactions. The pooled material was collected from donors prior to December 2019.

Serial dilutions of heat-inactivated SARS-CoV-2 viruses were done in VTM to concentrations ranging from 4×10^3^ copies/μL to 1 copy/μL in the starting sample. The samples were aliquoted into PCR tubes and lysed (95 °C, 1 min.) prior to adding RT-LAMP reaction mix. For our optimized 2-step protocol, the B3 and BIP primers and RT enzyme were added for the RT-incubation step (55°C, 10 min.). Remaining RT-LAMP reagents including F3, FIP and Loop primers were added for a final reaction volume of 16 μL. The amplification reaction was performed at 65°C for 60 min.

Reactions done with heat-inactivated SARS-CoV-2 viruses in saliva were in the same formats as mentioned above for the 1-step and our optimized 2-step RT-LAMP assays. Heat-inactivated SARS-CoV-2 viruses were serially diluted and directly spiked in saliva to starting sample concentrations ranging between 4×10^3^ copies/μL to 1 copy/μL. The samples were aliquoted into PCR tubes and thermally lysed at 95°C for 1 minute prior to adding the RT-LAMP reaction reagents. In our optimized 2-step RT-LAMP assay, 7.85 μL of RT reaction volume including the B3, BIP primers, RT enzyme, and buffer were added to 2 μl of sample for the RT-incubation step (55°C, 10 min). Remaining RT-LAMP reagents (6.15 μL) were added post RT step, for a final reaction volume of 16 μL. The amplification reaction was done in a thermocycler (65°C, 60 min.).

Prior to the thermal lysis step, the clinical samples (VTM and saliva) were aliquoted in two portions. One aliquot was used for internal control and therefore spiked with MS2 bacteriophage (ZeptoMetrix Corporation) in a 1:9 ratio (MS2:sample).

### Amplification Data Analysis

The RT-LAMP fluorescence curves and amplification threshold bar graphs were analyzed using a MATLAB script and plotted using GraphPad Prism 8. For each curve, the threshold time was taken as the time required for each curve to reach 20% of the total intensity. The amplification threshold bar graphs show the mean and SD of three samples for characterization experiments and the mean and SD of four samples for clinical samples analysis.

ROC analysis was performed on GraphPad Prism 8.

PPV = (Number of true positives)/(Number of true positives + Number of false positives).

NPV = (Number of true negatives)/(Number of true negatives + Number of false negatives).

Sensitivity = (Number of true positives)/(Number of true positives + Number of false negatives).

Specificity = (Number of true negatives)/(Number of true negatives + Number of false positives).

## Data Availability

All Data is included in the manuscript.

## Author Contributions

A.G. and R.B. designed the assay and diagnostics study. E.V, S.A.S.d.R, M.J., K.W., J.K., and R.B. designed the clinical sample collection protocols. A.G., A.M, J.B. performed the experiments. A.B., K.R., M.A., S.A., M.M., P.M., and S.S. supported the clinical study, collected samples and analysed data. A.G., A.M., J.B., E.V., and R.B. analysed data. A.G., A.M, E.V. R.B. wrote the manuscript.

## Acknowledgments

The following reagents were obtained through BEI Resources, NIAID, NIH: (i) Genomic RNA from Zika Virus, PRVABC59, NR-50244; (ii) Zika Virus, PRVABC59, NR-50240. The following reagents were deposited by the Centers for Disease Control and Prevention and obtained through BEI Resources, NIAID, NIH: (i) Genomic RNA from SARS-Related Coronavirus 2, Isolate USA-WA1/2020, NR-52285; (ii) SARS-Related Coronavirus 2, Isolate USA-WA1/2020, Heat Inactivated, NR-52286.

Authors thank the staff at the Holonyak Micro and Nanotechnology Laboratory at UIUC for facilitating the research and the funding from University of Illinois. R.B. and E.V. acknowledges support of NIH R21 AI146865A, and to also support A.G. A.M. was partially supported by a cooperative agreement with Purdue University and the Agricultural Research Service of the United States Department of Agriculture, AG Sub Purdue 8000074077 to R.B. R.B. and E.V. acknowledge the support of NSF Rapid Response Research (RAPID) grant (Award 2028431). R.B., E.V., A.G. and S.A.S.d.R acknowledge the support of Jump Applied Research through Community Health through Engineering and Simulation (ARCHES) endowment through the Health Care Engineering Systems Center at UIUC.

Authors thank Sara Riggenbach, Gabriel Koch, and Bill Bond of OSH Healthcare (Peoria, IL) for their support of the IRB # 1602513 and patient sample acquisition for this study. We also thank Mary Ellen Sherwood, Reubin McGuffin, and Carly Skadden of Carle Foundation Hospital (Urbana, IL) for their support of the IRB # 20CRU3150 and patient sample acquisition for this study.

## Supplementary Information

**Figure S1.**
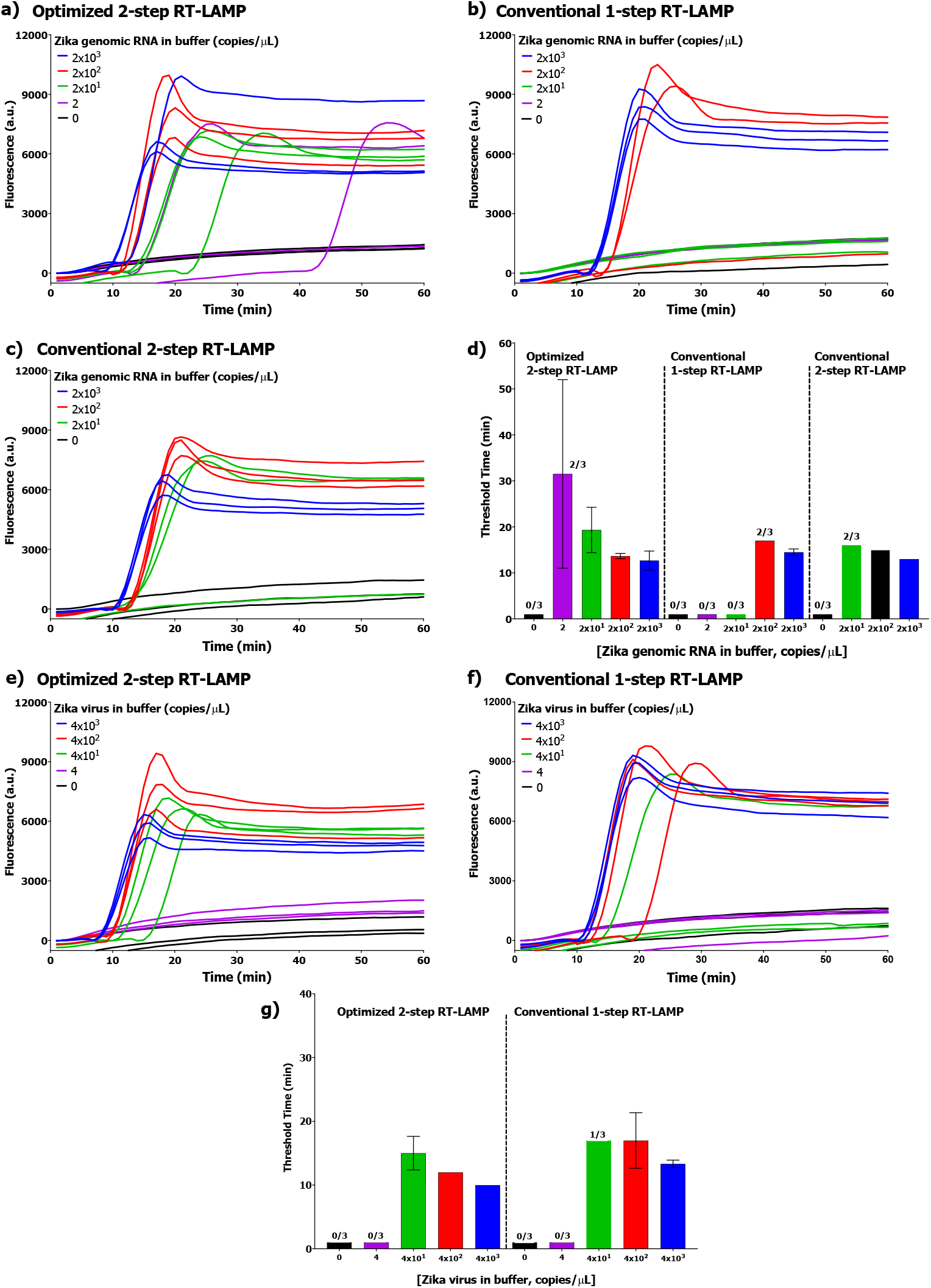
Detection of ZIKA genomic RNA and virus in buffer. **a-c)** Raw amplification fluorescence curves for Zika genomic RNA detection in buffer. **d)** Threshold timings for Zika genomic RNA detection in buffer. Optimized 2-step RT-LAMP LOD = 2 copies/μL; conventional 1-step RT-LAMP LOD = 2×10^3^ copies/μL; conventional 2-step RT-LAMP LOD = 2×10^2^ copies/μL. **e-f)** Raw amplification fluorescence curves for Zika virus detection in buffer; **g)** Threshold timings for Zika virus detection in buffer. Optimized 2-step RT-LAMP LOD = 4×10^1^ copies/μL; conventional 1-step RT-LAMP LOD = 4×10^2^ copies/μL. The bar graphs show mean and standard deviation for 3 replicates.

**Figure S2.**
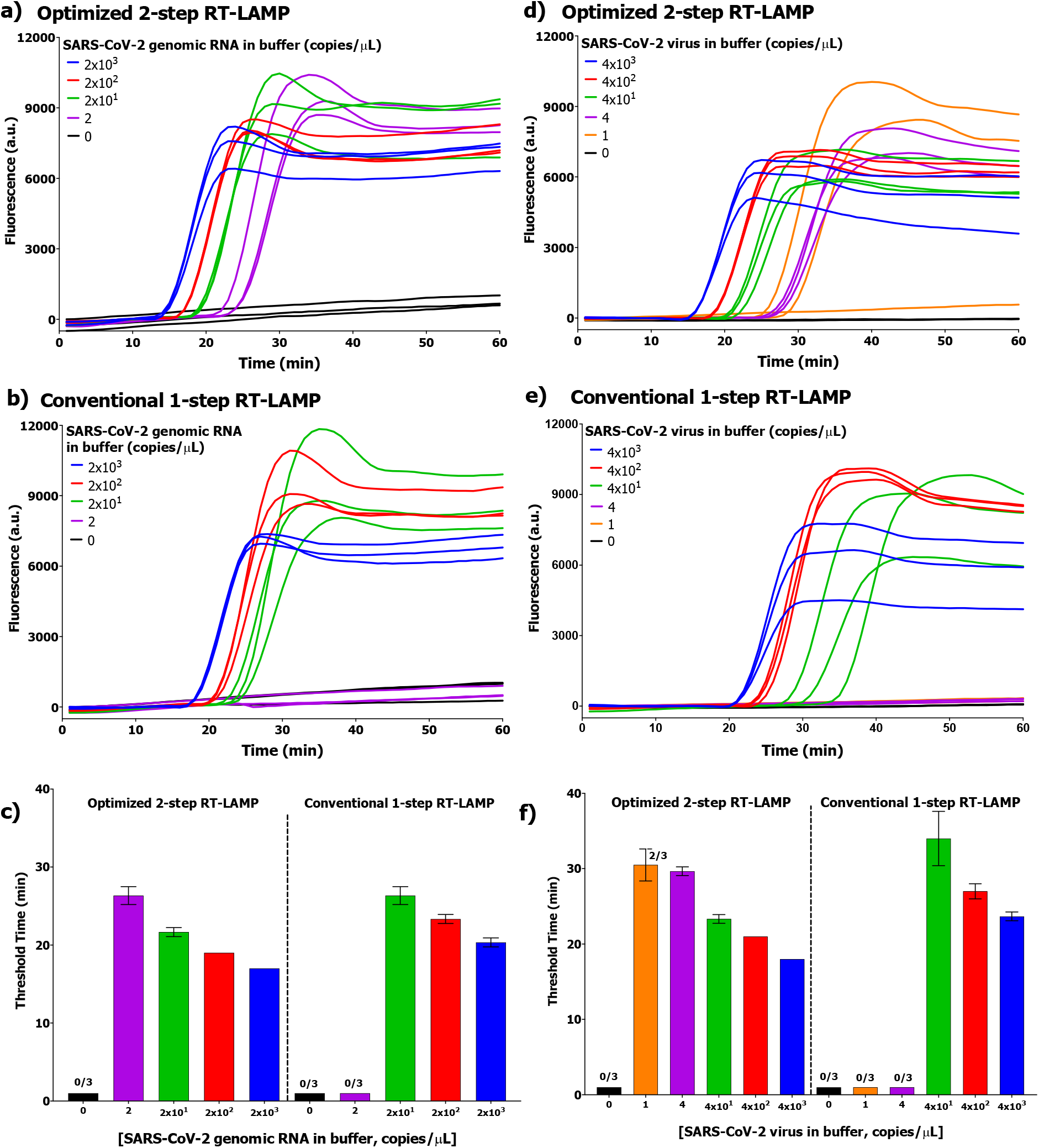
Detection of SARS-CoV-2 genomic RNA and virus in buffer. **a-b)** Raw amplification fluorescence curves for SARS-CoV-2 genomic RNA detection in buffer. **c)** Threshold timings for SARS-CoV-2 genomic RNA detection in buffer. Optimized 2-step RT-LAMP LOD = 2 copies/μL; Conventional 1-step RT-LAMP LOD = 2×10^1^ copies/μL. **d-e)** Raw amplification fluorescence curves for inactivated SARS-CoV-2 virus detection in buffer; **f)** Threshold timings for inactivated SARS-CoV-2 virus detection in buffer. Optimized 2-step RT-LAMP LOD = 1 copies/μL; Conventional 1-step RT-LAMP LOD = 4×10^1^ copies/μL. The bar graphs show mean and standard deviation for 3 replicates.

**Figure S3.**
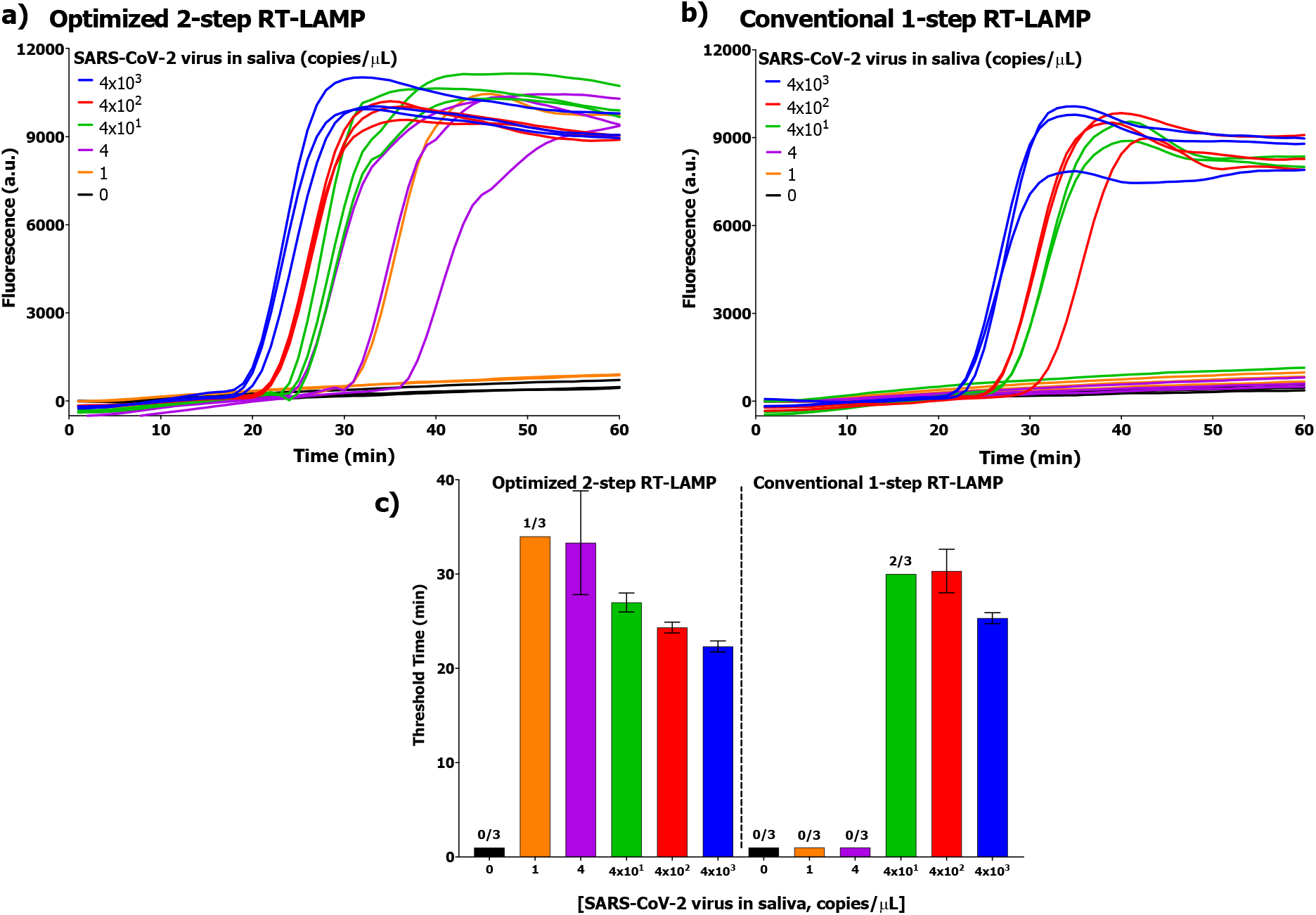
Detection of inactivated SARS-CoV-2 virus in human saliva (replicate data). **a-b)** Raw amplification fluorescence curves for inactivated SARS-CoV-2 virus detection in saliva; **c)** Threshold timings for inactivated SARS-CoV-2 virus detection in saliva. Optimized 2-step RT-LAMP LOD = 1 copies/μL; Conventional 1-step RT-LAMP LOD = 4×10^2^ copies/μL. The bar graphs show mean and standard deviation for 3 replicates.

**Figure S4.**
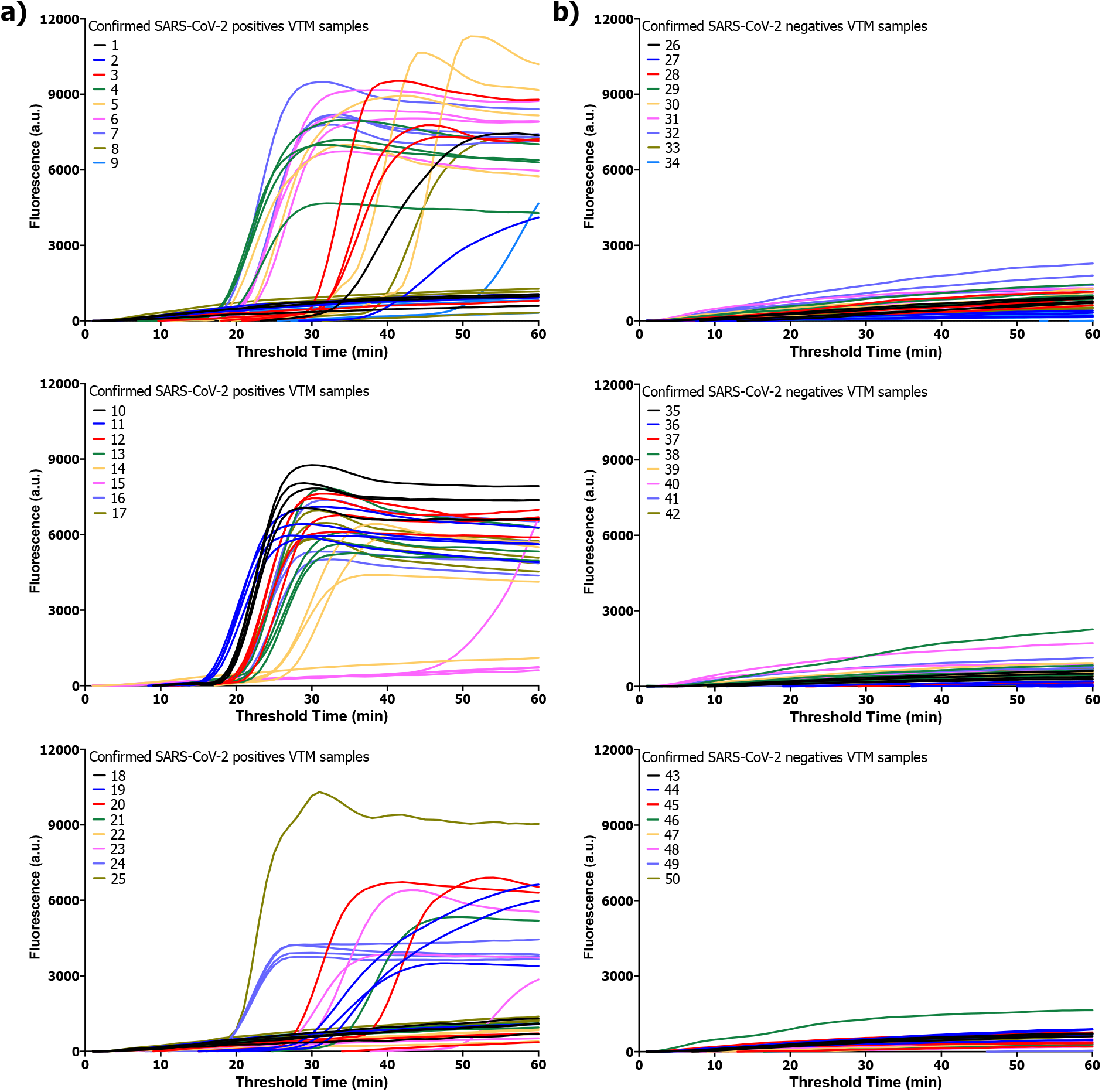
Amplification raw data of the results obtained when analyzing VTM clinical samples with our 2-step RT-LAMP reaction (small volume). **a)** Individual amplification curves of each confirmed positive sample (n = 4). **b)** Individual amplification curves of each confirmed negative sample (n = 4).

**Figure S5.**
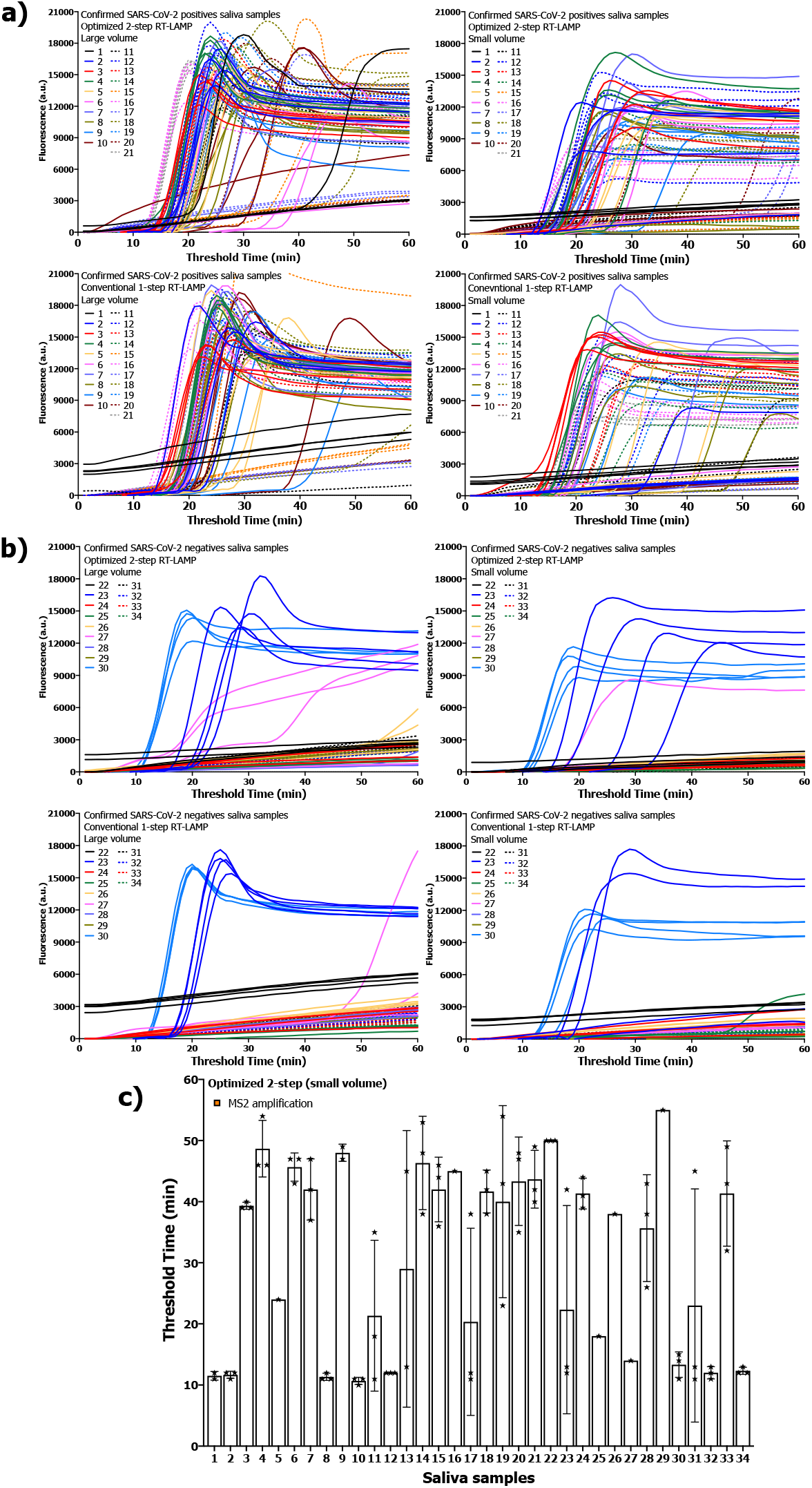
Saliva clinical samples results. **a)** Individual amplification curves of each confirmed positive sample (n = 4) in four different reaction formats: optimized 2-step RT-LAMP (large volume), optimized 2-step RT-LAMP (small volume), conventional 1-step RT-LAMP (large volume), and conventional 1-step RT-LAMP (small volume). **b)** Individual amplification curves of each confirmed negative sample (n = 4) in four different reaction formats: optimized 2-step RT-LAMP (large volume), optimized 2-step RT-LAMP (small volume), conventional 1-step RT-LAMP (large volume), and conventional 1-step RT-LAMP (small volume). For all saliva samples, internal control was also analyzed (n = 3) in the optimized 2-step RT-LAMP small volume format. Symbols in the bars indicate the results of the individual replicates.

